# A Systematic Review of the Current State of Trauma Clinical Practice Guidelines in the United States: Timeliness, Relevance, Accessibility, Quality, and Equity

**DOI:** 10.1101/2025.08.08.25333163

**Authors:** Maclean S. Panshin, Tina Samsamshariat, Gabriela Zavala Wong, Yahaira Carpio Colmenares, Ashley D. Farley, Sam Van Hook, Ariel W. Knight, Jakob E. Gamboa, Colby G. Simmons, Lacey N. LaGrone

## Abstract

**Background:** Clinical Practice Guidance (CPGs) are essential for standardizing trauma care and improving patient outcomes. Trauma clinical guidance in the United States often lacks coordination, timely updates, and applicability beyond academic Level I centers. This systematic review assesses the current state of trauma-related CPGs in the United States by examining their timeliness, relevance, accessibility, quality, and equity.

**Methods:** A systematic database search identified 1766 studies, of which 65 met inclusion criteria. We searched eight databases and 28 trauma societies’ websites for CPGs published between 2016 and 2024. We utilized MeSH terms to gather protocols, guidelines, and reviews, and evaluated their quality using the National Guideline Clearinghouse Extent Adherence to Trustworthy Standards (NEATS) criteria. Logistic regression was then used to analyze predictive factors for high-quality guideline creation.

**Results:** Most authors (96.7%) were affiliated with a Level I trauma center, with significant contributions from Trauma Region 3. Fifty-five (84.6%) CPGs were sponsored by professional societies, most commonly by the Eastern Association for the Surgery of Trauma (EAST). Eighty-three percent of included guidance was open access without requiring registration. The average cost of the remaining CPGs was 46.16 USD (SD 14.21). The mean NEATS score was 4.02 ± 1.01. Logistic regression showed that high-quality guidance was significantly associated with the inclusion of methodological experts (p=0.03) and funding disclosures (p=0.04).

**Conclusions:** Trauma CPGs are primarily produced by academic institutions affiliated with Level I trauma centers and are commonly endorsed by a professional trauma society. While most guidance is open access, there is an opportunity to enhance their quality, accessibility, and applicability, particularly in lower-resourced settings.

## BACKGROUND

Unintentional injury accounted for 218,064 deaths in the United States (US) in 2022 and remains the leading cause of death in individuals between 1 and 45 years old (1,2). Such high injury prevalence is compounded by substantial disparity in patient outcomes between trauma centers. Up to 18,000 trauma-related deaths may be prevented annually with widespread implementation of contemporary clinical management strategies (3). However, several barriers hinder access to trauma care. Geography, for example, contributes to disparities in trauma mortality, as rural patients experience nearly twice the rate of preventable trauma-related deaths compared to their urban counterparts. This difference is largely attributable to limited access to nearby Level I/II trauma centers, which are associated with improved outcomes (4,5).

A proposed strategy to standardize trauma care, improve outcomes across healthcare systems, and reduce preventable mortality is through the development, dissemination, and implementation of clinical practice guidance (CPGs). As defined by the Institute of Medicine (IOM), CPGs are “statements that include recommendations intended to optimize patient care that are informed by a systematic review of evidence and an assessment of the benefits and harms of alternative care options”(6). This guidance is developed by a range of entities, including clinical specialty societies, disease- and population-specific organizations, and individual medical centers. The IOM further recommends improved integration of clinical care and up-to-date science to support high-quality, evidence-based health systems (7). Effective CPGs are clearly written, focus on specific and relevant clinical problems, and are regularly updated to reflect the best available evidence (6). In the context of trauma care, the American College of Surgeons’ Committee on Trauma (COT) developed the Trauma Quality Improvement Program (TQIP) to enhance care quality by providing risk-adjusted benchmarking data to Level I and II trauma centers, along with targeted feedback to support improved clinical outcomes (8).

Given the substantial burden of injury in the United States, ensuring that high-quality trauma guidance is readily accessible to clinicians is essential for the delivery of optimal, evidence-based care. This systematic review evaluates the current landscape of national trauma guidance with respect to their timeliness, clinical relevance, accessibility, methodological quality, and equity considerations. In doing so, we aim to identify actionable opportunities to strengthen both the development and dissemination of CPGs in the United States.

## METHODS

This systematic review was conducted following the Preferred Reporting Items for Systematic Reviews and Meta-Analysis Protocols (PRISMA-P). The systematic review protocol was registered in the International Prospective Register of Systematic Reviews (PROSPERO) in accordance with the guidelines on 27 December 2021 (registration number CRD42021292381).

### Search Strategy

Two complementary search strategies were employed to identify relevant trauma CPG: a systematic keyword search of major electronic databases and a targeted review of trauma society websites.

The database search included PubMed, Embase, Google Scholar, SciELO, Global Index Medicus, EBSCO, ProQuest, and Ovid Medline. This comprehensive approach was designed to maximize the capture of published guidance across various disciplines and regions. The initial search was conducted in December 2021 and included guidance published between January 1, 2016, and December 5, 2021. A five-year window was selected to align with the median lifespan of clinical guidance. To ensure findings remained current, the search was updated in January 2024.

In parallel, a targeted review of the websites of key national and international trauma societies was performed to capture guidance that may not have been indexed in electronic databases. These included the American Association for the Surgery of Trauma (AAST); Eastern Association for the Surgery of Trauma (EAST); Western Trauma Association (WTA); International Association for Trauma Surgery and Intensive Care (IATSIC); Society for Critical Care Medicine (SCCM); American Trauma Society (ATS); World Congress of Abdominal Compartment Syndrome (WCACS); International Society of Surgery (ISS/SIC); Orthopaedic Trauma Association (OTA); and World Society of Emergency Surgery (WSES).

### Search Criteria

A keyword search of the databases was performed using the following terms: “trauma,” “burns,” “electrical injury,” “multiple trauma,” “traumatic brain injury,” “spinal cord injury,” “pelvic injury,” “urologic injury,” “concussion,” “contusion” “hemothorax” “pneumothorax,” “flail chest,” and “thoracic injury,” in combination with the Boolean operators “AND” and “OR.” Individual search strategies for each database can be found in the supplementary appendix.

### Inclusion Criteria

Source content was selected per the IOM CPG definition based upon the following criteria: establishment of transparency, management of conflicts of interest, multidisciplinary group composition, CPG-systematic review intersection, evidence-based foundations, rating strength of recommendations, and articulation of recommendations (9). Only guidance with at least one US author or US trauma society affiliation were included. CPGs and consensus statements that addressed any trauma-related clinical problem were included. Guidance addressing post-traumatic complications were included, as were dental and nursing guidance.

Prehospital studies, opinion pieces, and editorials were excluded. Abstracts not associated with a full-text publication were also excluded. No language-dependent exclusion criteria were applied, however, the keyword search was only conducted in English. Full texts not available in English or Spanish were translated into one of these languages to facilitate data abstraction.

### Data Extraction

Titles and abstracts identified through the literature search were first reviewed to remove duplicate records, then independently screened by three authors (M.S.P., T.S., and G.Z.W.) to assess potential eligibility. Each citation was evaluated by two authors, with any disagreements resolved by consensus with a third reviewer. Full-text analysis was subsequently conducted for studies meeting the inclusion criteria. The following information was abstracted from each study meeting the inclusion criteria: (a) date of publication, (b) whether website registration was required for access, (c) whether institutional membership provided access, (d) access cost, (e) number of clicks to access article, (f) clinical problem addressed, (g) authors’ originating trauma region, and (h) guideline quality. The number of clicks was determined by entering the article title into Google and counting the clicks needed to access the full text.

The quality of CPGs was assessed using the National Guideline Clearinghouse Extent Adherence to Trustworthy Standards (NEATS) instrument. Each criterion is rated on a scale from 1 (lowest) to 5 (highest), with scores greater than 4 indicating high.

### Data Analysis

Descriptive statistics related to author institutional affiliation, state, and region, as well as society contribution, accessibility, and quality. Logistic metrics were calculated and presented. Frequencies, means, and percentages are reported. Map charts were developed to visually depict the geographical distribution in guideline authorship and ACS trauma region. A logistic regression was used to identify factors performed to assess for associations between predictive factors and CPGs having a NEATS score greater than 4. A p-value of 0.05 was considered statistically significant. Analysis was conducted using Stata 18.0 (StataCorp 2023, College Station, TX) and Microsoft Excel.

## RESULTS

Our initial search yielded 1,766 studies (1,676 from the database search and 90 from professional trauma societies). After 780 duplicates were removed, 986 records were screened, and 948 met the inclusion criteria. After further assessment, 65 studies were included in the final systematic review sample (Figure 1).

**FIGURE 1.**
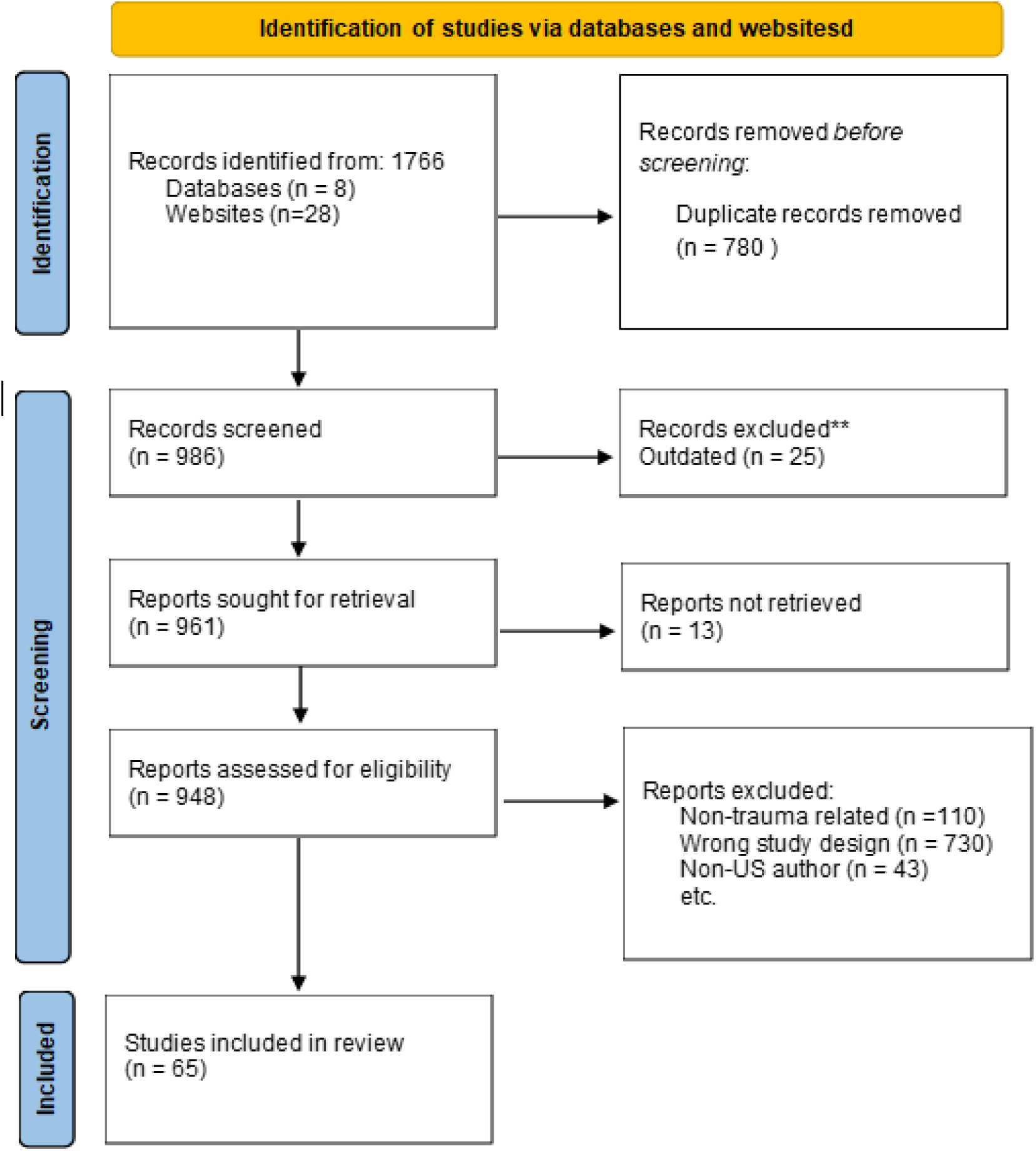
Study selection process.

Of the total author pool, 578 (96.7%) were affiliated with a Level I trauma center, yet only 31 first authors held a Level I affiliation, and only 18 (3%) authors were affiliated with a Level II trauma center. Geographically, 12.7% of authors were affiliated with institutions located in originated from California (n=84), followed by Pennsylvania (7.7%, n=51), Ohio (7.4%, n=49), and Washington (7.2%, n=48) (Figure 2a). Sixty-five guidance were produced by authors from the first 10 Trauma Regions designated by the American College of Surgeons Committee on Trauma (ACS COT). One guideline included authors from multiple trauma regions; these contributions are reflected in Trauma Regions (Figure 2b.). Overall, Trauma Region 3 (Delaware, District of Columbia, Maryland, Pennsylvania, West Virginia) contributed to the greatest number of guidance (n=56, 16.4%), followed by Region 9 (Arizona, California, Hawaii, Nevada) with 44 (12.9%). Region 4 (Alabama, Florida, Georgia, Kentucky, Mississippi, North Carolina, South Carolina, Tennessee), Region 5 (Illinois, Indiana, Michigan, Minnesota, Ohio, Wisconsin), and Region 10 (Alaska, Idaho, Oregon, Washington) contributed to 41 (12%), 40 (11.7%), and 34 (9.9%) guidance, respectively. Among corresponding authors, 8 of 37 (21.6%) were from Region 4. For first authors, 11 of 48 (22.9%) were based in Region 9. Region 3 accounted for 19 of 65 (29.2%) senior authors (Figure 2b).

**FIGURE 2.**
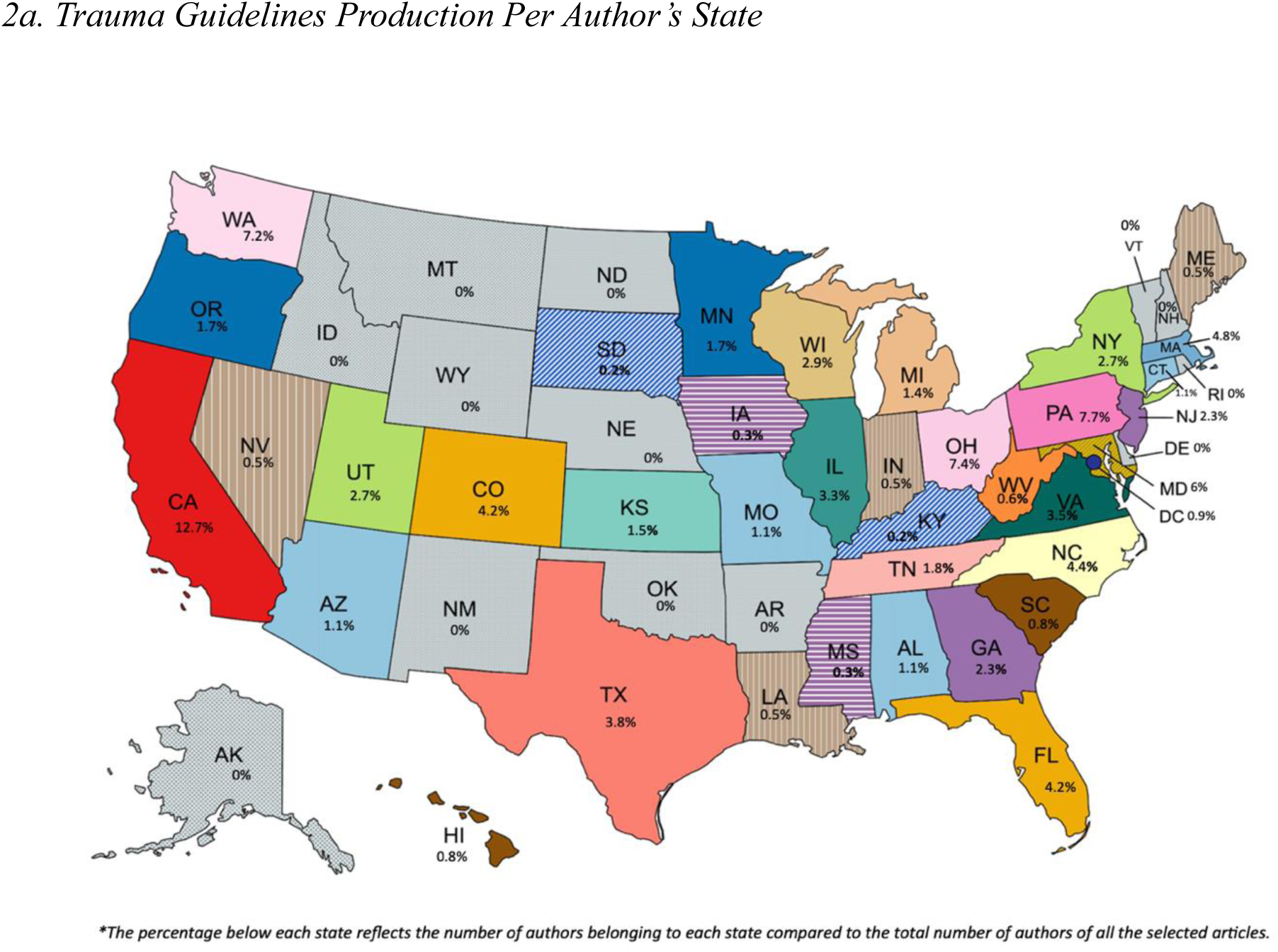
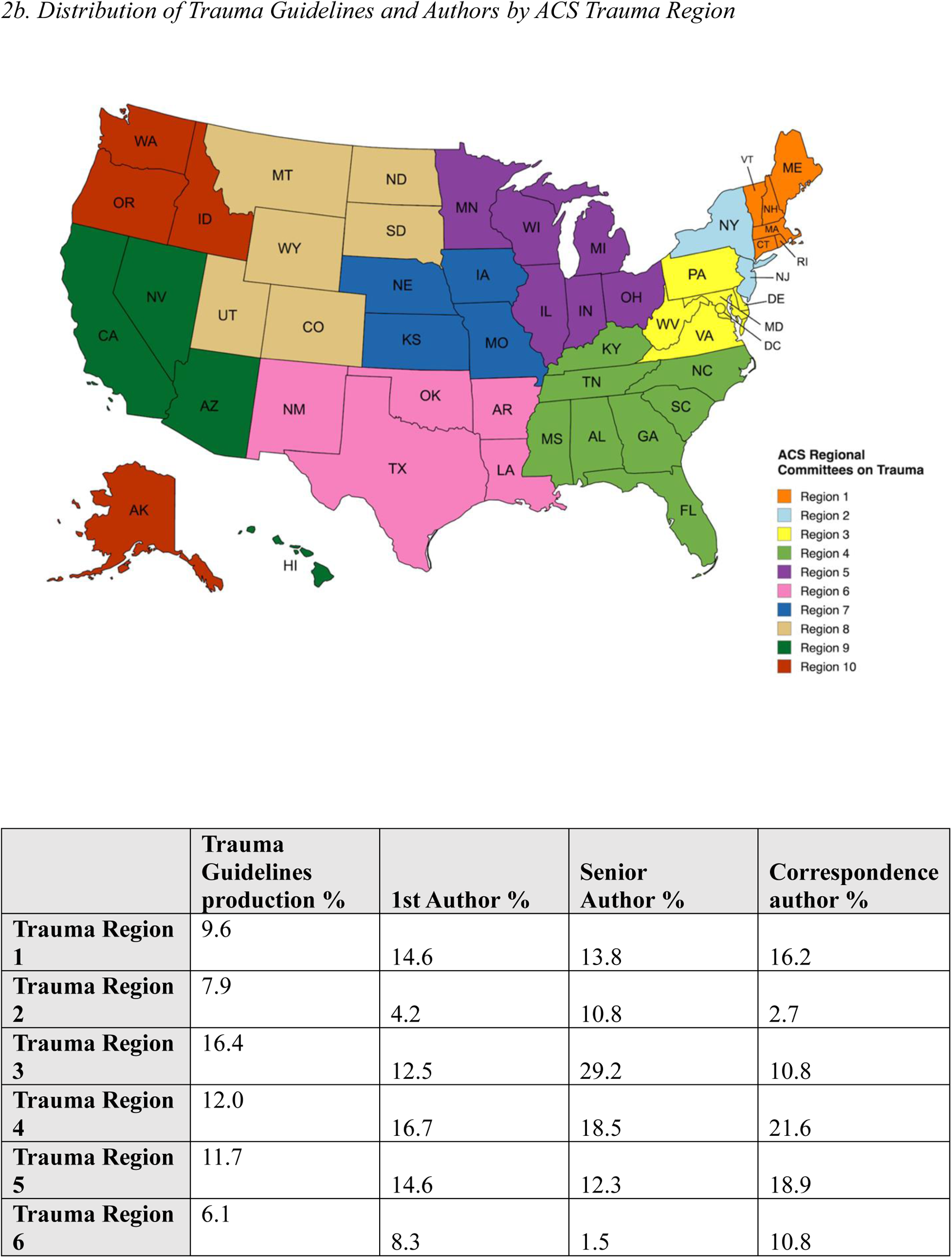
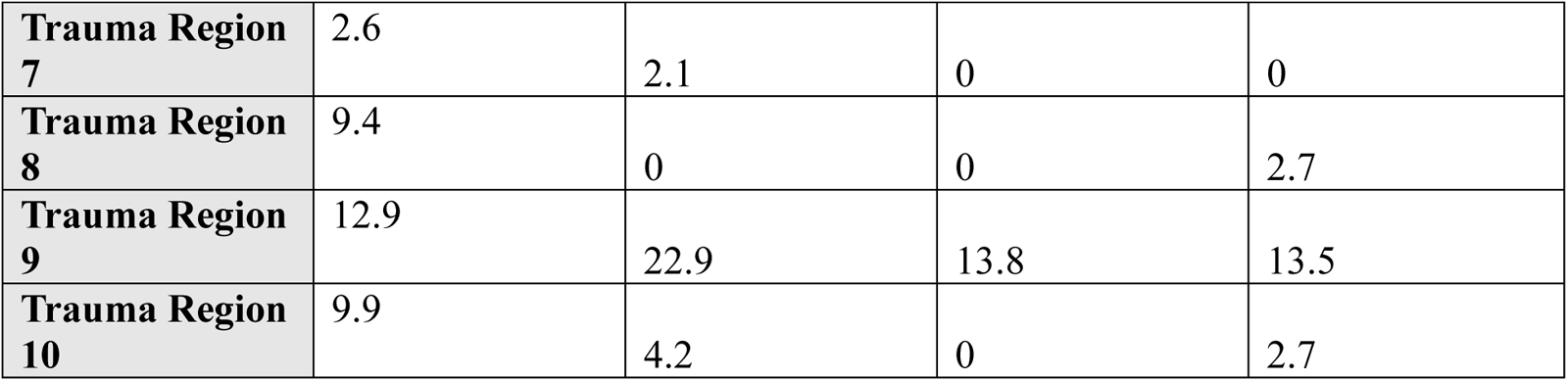
Trauma Guidelines Production.

The included guidance were sponsored by 16 distinct professional trauma societies. EAST produced the largest proportion, authoring 22 guidance (26.5%), followed by WSES with 18 (21.7%). All remaining societies contributed between one and four guidance each (Table 1). Regarding accessibility, 54 guidance were open access, defined as being available free of charge and without the need for institutional or professional society affiliation. In contrast, 8 guidance (12.4%) required society membership for access. For guidance behind a paywall, the mean cost to obtain full-text access was 46.16±14.21 US dollars (USD. On average, full-text guidance could be reached in two). Two or fewer clicks. All included guidance were published exclusively in English, and no issues were identified related to broken links or corrupted files (Table 2).

**TABLE 1.** Participation Of Society In Publications. ^*^*The number of publications in which each society appeared; some societies participated in the same article and were quantified separately.*

|  | **Number of publications\*** | **%** |
| --- | --- | --- |
| ***Eastern Association for the Surgery of Trauma (EAST)*** | 22 | 26.5% |
| ***World Society of Emergency Surgery (WSES)*** | 18 | 21.7% |
| ***Other societies (from other countries that are not US)*** | 17 | 20.5% |
| ***American Association and Congress of Neurological Surgeons*** | 4 | 4.8% |
| ***American Association for the Surgery for Trauma (AAST)*** | 4 | 4.8% |
| ***The AO Spine North America*** | 4 | 4.8% |
| ***American Academy of Orthopedic Surgeons (AAOS)*** | 2 | 2.4% |
| ***American Burn Association*** | 2 | 2.4% |
| ***Brain Trauma Foundation*** | 2 | 2.4% |
| ***American Academy of Physical Medicine and Rehabilitation*** | 1 | 1.2% |
| ***American Clinical Neurophysiology Society*** | 1 | 1.2% |
| ***Congress of Neurological Surgeons*** | 1 | 1.2% |
| ***Major Extremity Trauma and Rehabilitation Consortium*** | 1 | 1.2% |
| ***Neurocritical Care Society (NCS)*** | 1 | 1.2% |
| ***Pediatric Trauma Society*** | 1 | 1.2% |
| ***Trauma Anesthesiology Society*** | 1 | 1.2% |
| ***Trauma Update Network*** | 1 | 1.2% |

**TABLE 2.**
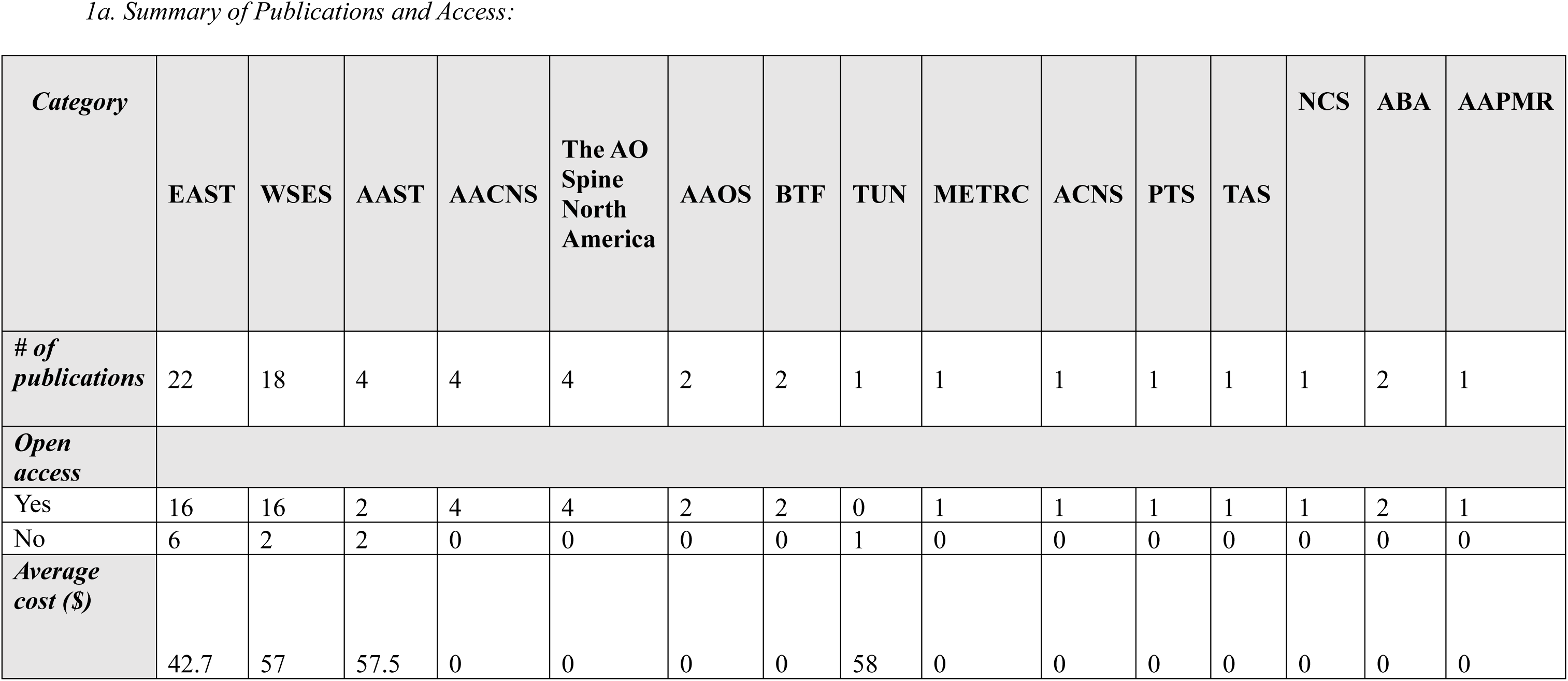
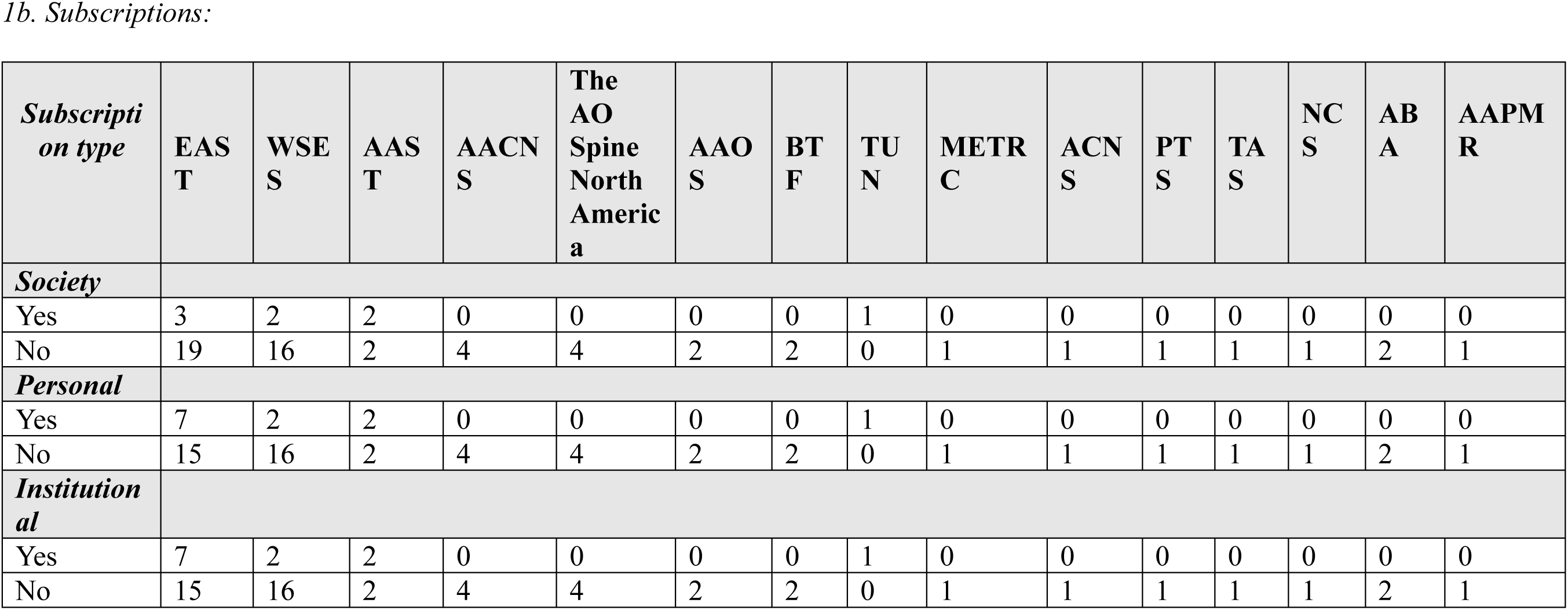
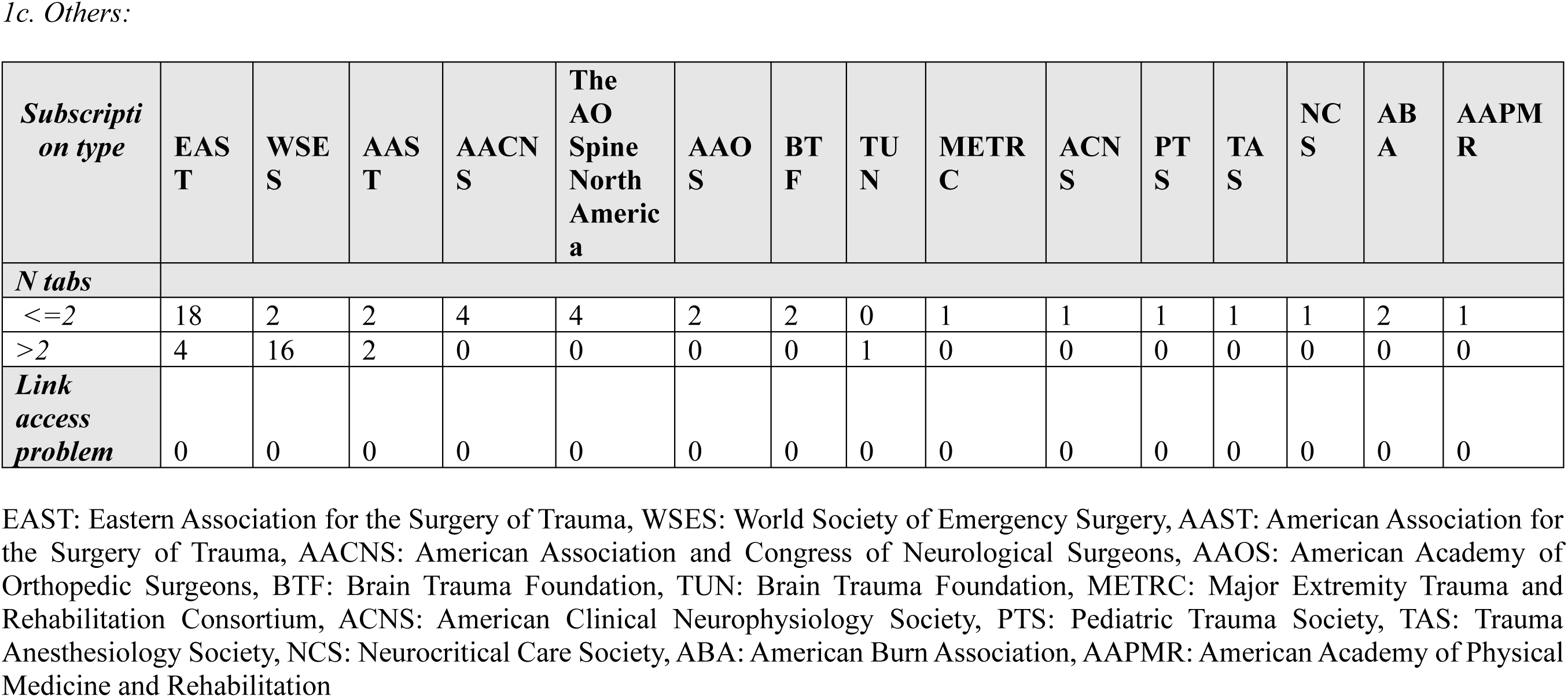
Access US paper by society.

All included guidance was assessed using the NEATS scorecard. Fifty (77%) guidance explicitly disclosed funding sources, and sixty-three guidance (97%) were developed by a multidisciplinary group. However, only 48 guidance (74%) reported involvement of a methodological expert. In logistic regression analysis, the inclusion of a methodological expert (p = 0.03) and disclosure of funding sources (p = 0.04) were both significantly associated with a high NEATS score (≥4) (Table 3).

**TABLE 3.** Quality Metrics based on NEATS scorecard.

|  | **2016**<br>**(N=5)** | **2017**<br>**(N=15)** | **2018**<br>**(N=4)** | **2019**<br>**(N=12)** | **2020**<br>**(N=9)** | **2021**<br>**(N=6)** | **2022**<br>**(N=9)** | **2023**<br>**(N=4)** | **2024**<br>**(N=1)** |
| --- | --- | --- | --- | --- | --- | --- | --- | --- | --- |
| **Disclosure of funding source<sup>¶</sup>** |  |  |  |  |  |  |  |  |  |
| Yes | 3 (60%) | 14 (93.3%) | 4 (100%) | 10 (83.3%) | 7 (77.8%) | 3 (50%) | 7 (77.8%) | 2 (50%) | 0 (0%) |
| No | 2 (40%) | 1 (6.7%) | 0 (0%) | 2 (16.7%) | 2 (22.2%) | 3 (50%) | 2 (22.2%) | 2 (50%) | 1 (100%) |
| **Presence of multidisciplinary group** |  |  |  |  |  |  |  |  |  |
| Yes | 5 (100%) | 15 (100%) | 4 (100%) | 12 (100%) | 8 (88.9%) | 6 (100%) | 8 (88.9%) | 4 (100%) | 1 (100%) |
| No | 0 (0%) | 0 (0%) | 0 (0%) | 0 (0%) | 1 (11.1%) | 0 (0%) | 1 (11.1%) | 0 (0%) | 0 (0%) |
| **Presence of a Methodologist<sup>¶</sup>** |  |  |  |  |  |  |  |  |  |
| Yes | 4 (80%) | 14 (93.3%) | 3 (75%) | 10 (83.3%) | 3 (33.4%) | 4 (66.7%) | 5 (55.6%) | 4 (100%) | 1 (100%) |
| No | 1 (20%) | 1 (6.7%) | 1 (0%) | 2 (16.7%) | 6 (66.6%) | 2 (33.3%) | 4 (44.4%) | 0 (0%) | 0 (0%) |
| **Disclosure of conflicts of interest\*** | 4.80 | 4.66 | 5 | 4.58 | 4.33 | 4 | 4.22 | 4 | 1 |
| **Patient and Public Perspectives\*** | 1.80 | 2.93 | 2.50 | 2.50 | 2.22 | 3.50 | 2.11 | 2 | 1 |
| **Use of a systematic review\*** | 4.60 | 4.53 | 5 | 4.16 | 4.55 | 3.66 | 4.77 | 5 | 5 |
| **Grading the quality of evidence\*** | 4.40 | 5 | 5 | 4.58 | 4.88 | 4.16 | 3.88 | 4.50 | 5 |
| **Benefits and harms of recommendations\*** | 4.40 | 4.46 | 4.75 | 4.58 | 4.55 | 4.33 | 4.11 | 4.75 | 5 |
| **Evidence supporting recommendations\*** | 4.20 | 4.73 | 4.75 | 4.66 | 4.77 | 4.66 | 4.55 | 4.50 | 5 |
| **Rating the strength of recommendations\*** | 4.60 | 4.93 | 5 | 4.58 | 4.55 | 4.16 | 4.33 | 4.75 | 5 |
| **Specific and Unambiguous Articulation of Recommendations\*** | 4.40 | 4.60 | 4.75 | 4.66 | 4.66 | 4.66 | 4.77 | 4.75 | 5 |
| **External review\***<br>**(scientific and clinical experts, organizations, patients, and representatives of the public)** | 3.60 | 4.33 | 4 | 4 | 4.22 | 4.16 | 3.88 | 3.75 | 3 |
| **Updating plans\*** | 1 | 2.53 | 1.50 | 1.58 | 1.22 | 1.33 | 2.77 | 1.4 | 1 |

\*This reflects an average
¶ $p < 0.05$

## DISCUSSION

This systematic review characterizes the development of trauma CPGs in the United States over the past eight years. CPGs are widely regarded as the gold standard for guiding clinical decision-making to optimize patient care and improve clinical outcomes (10). Trauma-specific CPGs aim to optimize the care of injured patients, disseminate evidence-based clinical recommendations, and enhance the efficiency of institutions involved in trauma care (11).

Among the 65 CPGs included in this review, Trauma Region 3 demonstrated the highest level of authorship participation, contributing to 56 guidance. This region encompasses Pennsylvania and Maryland, home to several nationally recognized trauma centers affiliated with surgical residency and fellowship programs, including Penn Presbyterian Medical Center (PPMC), Temple University Hospital (TUH), the R Adams Cowley Shock Trauma Center at the University of Maryland, and Prince George’s Hospital Center (PGH). These institutions are distinguished by a high volume of trauma-related clinical activity and sustained scholarly output. For example, the Shock Trauma Center manages approximately 8,000 critically ill and severely injured patients annually and is widely regarded as an international leader in trauma care and research (12). PPMC integrates research, education, and care through its robust residency and fellowship programs and the PennSTAR flight program (13). PGH, a Level II Trauma Center, treats a high volume of penetrating trauma cases and actively contributes to research and surgical training efforts through its affiliation with the University of Maryland (14). The combination of high clinical volume and robust research infrastructure. Likewise, California was the state with the highest contribution of CPG authors, likely due to the large number of trauma centers and academic institutions. The high level of clinical and academic activity at these institutions likely positions them as national leaders in the development of trauma clinical practice guidance.

The majority of authors (96.7%) were affiliated with a Level I trauma center. Level I trauma centers provide the highest level of comprehensive, multidisciplinary care, including trauma prevention and rehabilitation. Surgeons at these centers are also frequently involved in surgical education and research (15,16). Additionally, Level I centers are more likely to sustain dedicated trauma research programs, supported by greater personnel and financial resources (16). For example, in California (Region 9), most trauma research is conducted at academically affiliated Level I/II trauma centers (19).

Furthermore, trauma surgeons affiliated with academic Level I trauma centers are more likely to belong to and participate in national trauma societies, which often serve as key platforms for research dissemination and CPG development (16). Reflecting this, most included CPGs (84.6%) were sponsored by a national trauma society. As of 2020, EAST has published more than 70 CPGs, collectively generating approximately 400 citations (20). In the present review, EAST was the most frequently represented organization, contributing to 22 guidance (33.8%).

Encouragingly, 75% of included CPGs were publicly accessible without requiring society membership or a paid journal subscription. This ease of accessibility is particularly important in trauma and acute care surgery, where time-sensitive clinical decisions necessitate immediate access to evidence-based guidance. Open access to CPGs not only supports high-quality, efficient care in emergent settings but also promotes health equity by reducing barriers for clinicians practicing in resource-limited or underserved environments. Conversely, the remaining 25% of guidance that require payment or affiliation to access highlights an ongoing opportunity to broaden access and ensure that critical clinical guidance is universally available.

Analysis of NEATS scorecard data highlights opportunities to optimize the development of trauma CPGs. Logistic regression analysis identified the inclusion of a methodological expert as an independent predictor of higher NEATS scores, yet over 25% of multidisciplinary guideline panels did not report such involvement. Incorporating methodological expertise is likely to enhance the rigor of CPG development through improved planning, execution, and data interpretation (21). Higher NEATS scores were also independently associated with disclosure of funding sources, which may reflect increased transparency and perceived credibility of guideline development processes (22).

Regional disparities in authorship also warrant attention. As demonstrated in Figure 2b, guideline development was heavily concentrated in a small number of trauma regions, while others, such as Regions 7 and 8, had minimal representation. This imbalance may contribute to geographic inequities in guideline dissemination and implementation. Increasing the engagement of trauma centers across all regions, especially those historically underrepresented in national guideline development, is a critical step toward ensuring that clinical practice guidance reflects diverse care environments.

### Limitations

This review has several limitations. Not all variables of interest were consistently reported across the included CPGs, and both the sample size and study period were relatively limited. Furthermore, there is a paucity of data evaluating the impact of CPG adherence on patient outcomes, particularly in lower-level trauma centers and resource-limited settings. Given that most CPGs are developed at academic Level I trauma centers, recommendations may not be feasibly implemented in differently resourced clinical environments. Notably, only 3% of authors were affiliated with Level II trauma centers. Expanding authorship to include a broader range of institutions, including non-academic and lower-resourced settings, may enhance the applicability and contextual relevance of future guidance.

### Conclusions

In summary, trauma CPGs in the United States predominantly originate from Level I trauma centers and are often developed through the efforts of multidisciplinary teams affiliated with national trauma societies. These institutions benefit from strong academic infrastructures and substantial research resources, which support both the development and dissemination of clinical guidance. While quality may be improved through increased methodological input and transparency in funding, future efforts must also address persistent equity gaps. Enhancing open access, broadening geographic and institutional authorship, and ensuring the feasibility of implementation across diverse healthcare settings will be essential to improving the reach and impact of trauma clinical guidance nationwide. Engaging underrepresented regions and non-academic trauma centers in guideline development may help ensure that future CPGs are more contextually relevant and broadly applicable across diverse care settings.

## Data Availability

All data produced in the present study are available upon reasonable request to the authors

